# Meta-analysis comparing the effect of combined acupuncture plus statin therapy versus statin therapy alone on lipid levels of patients with angina pectoris of coronary heart disease

**DOI:** 10.1101/2024.02.14.24302641

**Authors:** Zhaobo Yan, Mailan Liu, Xianming Wu, Jiaojiao Xiong, Zhihong Yang, Ning Zhang, Xiaofang Yang, Mi Liu

## Abstract

**Objectives:** Establish clinical evidence regarding acupuncture combined with statin for blood lipid control in patients with angina pectoris (AP) of coronary heart disease (CHD) by systematically analyzing data from all available studies.

**Design:** A systematic review and meta-analysis.

**Data sources:** The literature search involved eight databases (China National Knowledge Infrastructure, Chinese Biomedical Literature Database, Wanfang, VIP Database for Chinese Technical Periodicals, PubMed, Embase, Web of Science, and Cochrane Library) and was finished on February 1, 2024.

**Eligibility criteria:** Random controlled trials (RCTs) investigating the efficacy of acupuncture combined with statin on lipid levels in patients with AP of CHD were eligible. The outcomes assessed were the lipid metabolism indicators, including TC, TG, LDL-C, and HDL-C, as well as the safety of the treatment.

**Data extraction and synthesis:** Data extraction and quality evaluation were conducted by two independent reviewers, with any discrepancies discussed by a third researcher. Pooled mean differences (MD) with 95% confidence intervals (CIs) were calculated for each outcome. Sensitivity and subgroup analyses were carried out to explore the heterogeneity. Publication bias was assessed using a funnel plot. The quality of the evidence was evaluated using the GRADE system.

**Results:** The final meta-analysis included nine eligible studies involving 754 patients. When comparing statin group, the acupuncture plus statin group showed lower levels of TC (MD=-0.48, 95% CI: −0.61 to −0.35, *P*<0.00001), TG (MD=-0.59, 95% CI: −0.86 to −0.32, *P*<0.00001), LDL-C (MD=-0.66, 95% CI: −0.99 to −0.33, *P*=0.0001), and higher levels of HDL-C (MD=0.16, 95% CI: 0.06 to 0.26, *P*=0.001). Each study included in the analysis exhibited some degree of bias. Significant publication bias was detected for the primary outcomes. Evidence quality for the primary outcomes was graded as very low.

**Conclusions:** Acupuncture as an adjunctive treatment can further improve lipid profile in individuals diagnosed with AP of CHD based on statin therapy. However, the clinical significance of this effect remains unclear; it is necessary to confirm the findings through more high-quality RCTs in the future.

**Trial registration number:** PROSPERO CRD42023465292.

**STRENGTHS AND LIMITATIONS OF THIS STUDY:**

1. This comprehensive systematic review will collect data regarding the combined therapy (acupuncture + statin) on lipid profiles in patients with AP of CHD to provide new evidence supporting the use of acupuncture treatment for AP of CHD.
2. Subgroup analysis based on the duration of intervention and types of acupuncture will provide a more detailed evaluation of the effectiveness of the combined therapy.
3. A thorough evaluation of the quality of the evidence for the primary outcomes was conducted using the Grading of Recommendation Assessment, Development, and Evaluations (GRADE) system.
4. This review has limited ability to draw clear conclusions due to poor quality of evidence.
5. We restricted the literature search to eight electronic databases and ignored a search for gray literature, which may miss important literature and could introduce publication bias.

## 1 Introduction

Coronary heart disease (CHD) is a prevalent cardiovascular disease and a vital cause of death worldwide.^1^ The primary pathophysiological basis for CHD is the development of atherosclerotic lesions in coronary artery vessels that can lead to narrowing or obstruction of the coronary lumen and predispose individuals to clinical manifestations related to myocardial ischemia and hypoxia, such as angina pectoris (AP).^2^ AP is the most common symptom experienced by CHD patients, characterized by chest pain, tightness, emotional upset, and shortness of breath.^3^ Given its significant impact on CHD patients’ quality of life and mortality, AP has become a growing public health concern.^4^ Dyslipidemia, an independent risk factor for AP of CHD, is a key mediator of the formation of atherosclerotic plaques, playing a crucial role in the progression of AP of CHD.^5^ ^6^ Furthermore, evidence suggests that individuals with elevated blood cholesterol, especially low-density lipoprotein (LDL), are at a higher risk of developing CHD and more susceptible to cardiovascular events.^7^ ^8^ Therefore, lipid-lowering therapy has become a key component in CHD treatment.

Statins are widely used for lipid management in patients with AP, as they have been proven effective in reducing the risk of cardiovascular events and mortality by controlling cholesterol levels.^9^ ^10^ Despite the efficacy of statins, some patients may not be able to tolerate high-intensity statin therapy or may not experience sufficient cholesterol reduction from statin therapy alone.^11^ ^12^ To help more patients achieve their lipid-lowering goals, the combination therapy of maximally tolerated statin with another non-statin lipid-lowering agent has been recommended by current guidelines.^9^ ^13^ ^14^

Acupuncture is a commonly complementary therapy that achieves therapeutic effects by stimulating some points in the body.^15^ It has beneficial effects on the management of AP associated with CHD. Several meta-analyses have demonstrated the effectiveness of acupuncture in reducing angina attack frequency and angina pain intensity and improving negative mood.^16–18^ Interestingly, extensive research has confirmed that stimulation over specific acupoints also has a beneficial impact on lipid metabolism, and the underlying mechanism for this effect may be associated with reducing liver lipid deposition, improving insulin resistance, and regulating glycerophospholipid and sphingolipid metabolism.^19–21^ Nowadays, there is increasing evidence suggesting that acupuncture treatment can improve lipid levels in patients with AP of CHD and has a synergistic effect when combined with a statin.^22–24^ However, relevant evidence-based medical research to guide clinical practice still does not exist. Therefore, this study aims to evaluate the hypolipidemic effect and safety of combining acupuncture and statin therapy in patients with AP of CHD by conducting a comprehensive meta-analysis of all available clinical evidence.

## 2 METHODS

This meta-analysis adhered to the guidelines of the Cochrane Handbook for Systematic Reviews of Interventions, and we checked the process of this review using the PRISMA (Preferred Reporting Items for Systematic Reviews and Meta-Analysis) checklist (Online supplementary Table 1).^25^ We registered this systematic review on the PROSPERO website with registration number CRD42023465292.

### 2.1 Data sources and search strategy

We conducted a literature search on acupuncture for AP of CHD, using eight electronic data sources: China National Knowledge Infrastructure (CNKI), Chinese Biomedical Literature Database (CBM), Wanfang, VIP Database for Chinese Technical Periodicals, PubMed, Embase, Web of Science, and Cochrane Library. We conducted this search process on February 1, 2024, with no limitations on language, geography, or year of publication. However, we only included literature published in Chinese or English for analysis. Two independent reviewers (Zhao-bo Yan and Xian-ming Wu) completed the retrieval process.

Our search strategy included three sets of terms: one focused on angina and CHD, another on lipid profile, and a third on acupuncture techniques. The search terms within each topic group were connected using the Boolean operator ‘OR,’ and then the three groups were linked using the ‘AND’ operator. The complete search strategy used in the PubMed database is shown in Table 2. The search terms were adjusted as necessary for other databases. To organize all the references obtained from the databases, we utilized EndNote (Version X. 9).

**Table 1.**
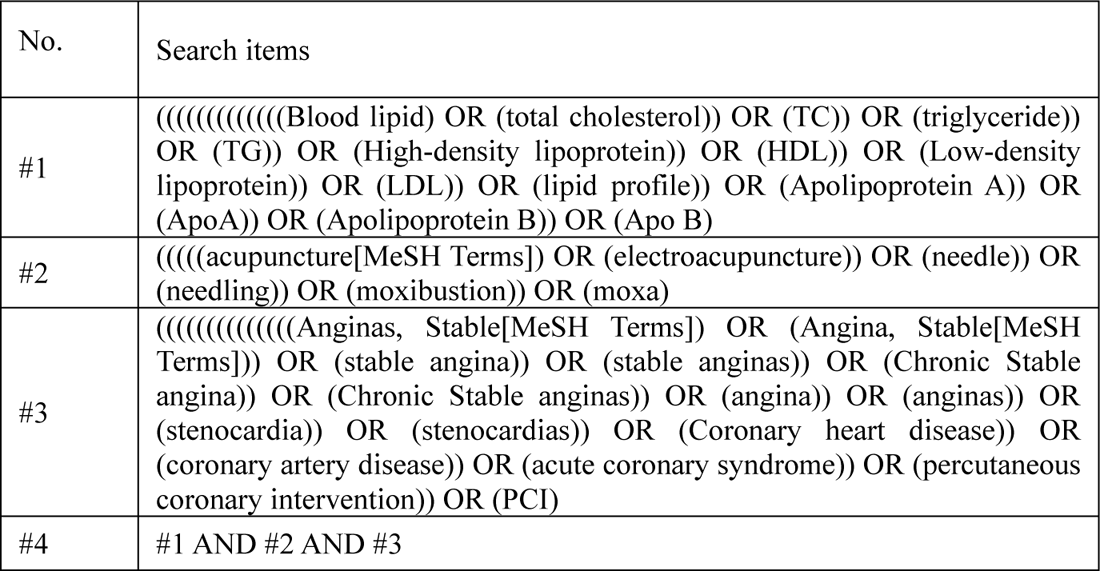
Search strategy in PubMed.

**Table 2.** Characteristics of the included studies.

| Study ID | Cases<br>(T/C) | Age<br>(Mean±SD, year) | Intervention |  | Statin prescription | Acupoints | Frequency and duration | outcomes |
| --- | --- | --- | --- | --- | --- | --- | --- | --- |
|  |  |  | T | C |  |  |  |  |
| Deng 2021 [29] | 34/34 | T: 50.87±10.23<br>C: 52.94±11.52 | Manual acupuncture +<br>Atorvastatin calcium tablets | Atorvastatin<br>calcium tables | oral, 20mg per<br>day for 4 weeks. | Main acupoints: CV17, BL15, PC6, ST36, SP6.<br>Cold coagulation and blood stasis syndrome:<br>BL14, CV4;<br>Qi deficiency and blood stasis syndrome: CV6;<br>Phlegm and blood stasis syndrome: ST40;<br>Qi stagnation and blood stasis syndrome: LR3. | 30-minute sessions once a day for<br>4 weeks. | TC, HDL-C,<br>TG, LDL-C |
| Jia 2016 [30] | 30/30 | Unknow | Electroacupuncture<br>(continuous-wave, 15 times/min)<br>+<br>Atorvastatin calcium tablets | Atorvastatin<br>calcium tables | oral, 10mg per<br>day for 4 weeks. | PC6 | 20-minute sessions once a day for<br>4 weeks. | TC, HDL-C,<br>TG, LDL-C,<br>Adverse event |
| Liu 2015 [22] | 48/48 | T: 52.96±9.05<br>C: 50.89±9.72 | Moxibustion +<br>Pravastatin sodium tablets | Pravastatin<br>sodium tablets | oral, 20mg per<br>day for 4 weeks. | BL15, BL14, CV17, PC6, SP6 | 10~40min per acupoint, once a<br>day, for 4weeks. | TC, HDL-C,<br>TG, LDL-C |
| Liu 2021 [31] | 30/30 | T: 54.79±3.92<br>C: 55.03±3.11 | Moxibustion +<br>Atorvastatin calcium tablets | Atorvastatin<br>calcium tables | oral, 20mg per<br>day for 8 weeks. | BL14, BL15, CV17, PC6, BL17. | 10~50min per acupoint, once a<br>day, for 8 weeks. | TC, HDL-C,<br>TG, LDL-C,<br>Adverse event |
| Sun 2021 [23] | 58/58 | T: 60.89±2.45<br>C: 60.62±3.47 | Manual acupuncture +<br>Atorvastatin calcium tablets | Atorvastatin<br>calcium tables | oral, 20mg per<br>day for 12 weeks. | HT7, PC6, ST40, SP9, CV6. | 30-minute sessions every 2 days<br>for 12 weeks. | TC, TG,<br>LDL-C |
| Wang 2015 [33] | 50/50 | T: 67±1<br>C: 67±2 | Moxibustion +<br>Atorvastatin calcium tablets | Atorvastatin<br>calcium tables | oral, 10mg per<br>day for 4 weeks. | PC4, HT6, CV14, CV17, BL17. | 30-minute sessions once a day,<br>continue 10 days for one<br>treatment course. This course of<br>treatment was repeated for a total<br>of 3 times, with a 2-day break<br>between each course. | TC, HDL-C,<br>TG, LDL-C |
| Wang 2021 [24] | 64/63 | T: 50±10<br>C: 49±10 | Manual acupuncture +<br>Atorvastatin calcium tablets | Atorvastatin<br>calcium tables | oral, 20mg per<br>day for 8 weeks. | PC5, LI11, PC6, PC7, HT7, LU9, KI3, BL66,<br>CV14, BL15. | 20-minute sessions three times a<br>week for 8 weeks. | TC, TG |
| Xue 2012 [32] | 34/33 | T: ≥50, n=22;<br><50, n=12.<br>C: ≥50, n=20;<br><50, n=13. | Manual acupuncture +<br>Simvastatin tablets | Simvastatin tablets | oral, 20mg per<br>day for 24 weeks. | BL15, PC6, BL14, CV17, CV26, HT7, LR3,<br>LR2, LR4, SP10, BL17, SP6. | 20-minute sessions once a day for<br>24 weeks. | TC, HDL-C,<br>TG, LDL-C |
| Zhao 2023 [34] | 30/30 | T: 54.46±9.16<br>C: 55.18±8.57 | Electroacupuncture<br>(continuous-wave, 15 times/min)<br>+<br>Atorvastatin calcium tablets | Atorvastatin<br>calcium tables | oral, 20mg per<br>day for 4 weeks. | Main acupoints: three special acupoints in cheek,<br>PC6, CV17, CV6, SP10, HT6, PC4;<br>Add-on BL15, BL17 for patients with blood stasis<br>syndrome. | 30-minute sessions once time, 3<br>times a week, for 4 weeks. | TC, HDL-C,<br>TG, LDL-C |

### 2.2 Eligibility criteria

#### 2.2.1 Types of studies

This study only included randomized controlled trials (RCTs) that assessed the effectiveness and safety of acupuncture for AP of CHD. Non-RCT research, such as single-arm trials, cohort studies, case reports, animal studies, qualitative studies, and conference abstracts, were excluded. Additionally, studies with missing or incomplete data were also excluded.

#### 2.2.2 Types of participants

This study included patients of any age, gender, race, or nationality diagnosed with AP of CHD according to published guidelines or criteria.

#### 2.2.3 Types of interventions

Clinical studies were eligible if the control group received statin alone while the intervention group received a combination of acupuncture and statin. We identified acupuncture as the treatment method stimulating acupoints by needle insertion or ignited moxa. Other AP of CHD medications, including nitroglycerin, aspirin, or trimetazidine, were allowed to be administered to both groups in the same way. Studies were excluded if the intervention group received other complementary or alternative therapies apart from acupuncture or if the control group received different types of acupuncture.

#### 2.2.4 Types of outcome measures

The clinical studies were eligible if they provided data on lipid profiles such as TC, TG, LDL-C, and HDL-C and information on adverse effects (AEs).

### 2.3 Study selection and data extraction

This procedure involved the participation of two independent reviewers, Zhao-bo Yan and Xian-ming Wu. Initially, duplicate articles were removed from the search results using Endnote (X9). Next, the titles and abstracts of the remaining literature were screened to identify irrelevant studies for further exclusion. After completing the above steps, we accessed the full text of the remaining articles and examined them to determine their eligibility for the final review. The vital information from the eligible studies, such as the first author, publication year, country, age, sample size, interventions, and outcomes, was recorded on a data extraction sheet. In the next step, two independent reviewers cross-checked the extracted data, resolving any inconsistencies through discussions and consultation with an experienced researcher, Xiao-fang Yang. In cases where the text contained unclear information, we would contact the corresponding author via email to obtain further details.

### 2.4 Quality assessment

Two independent reviewers (Zhao-bo Yan and Xian-ming Wu) conducted the methodological quality assessment for each included study using the “risk of bias” tool of Cochrane Collaboration.^26^ Six risks of bias domains of this tool were all applied in the present review: selection bias, performance bias, detection bias, attrition bias, reporting bias, and other bias. Based on the Cochrane Handbook, each study was considered a High, Low, or Unclear risk of bias in each domain. A third researcher (Xiang-fang Yang) adjudicated any discrepancies between the two reviewers.

### 2.5 Quality of evidence

We used the GRADE Pro GDT application to evaluate the quality of evidence for the primary outcomes.^27^ We assessed the impact of factors such as risk of bias, consistency, precision of effects, publication bias, and indirectness of evidence on the quality of evidence, following GRADE quality assessment criteria. Finally, the GRADE system classified the evidence as ‘high,’ ‘moderate,’ ‘low,’ or ‘very low’ quality. We involved three reviewers in this process (Zhao-bo Yan, Xian-ming Wu, and Xiao-fang Yang).

### 2.6 Data synthesis and analysis

All data were pooled using Review Manager Software (5.3 version). Because all blood lipid data were reported in the same format (mean ± standard) and used the same units (mmol/L), we calculated the weight mean differences (WMD) with 95% confidence intervals (CIs) to assess group differences in lipid levels. According to Cohen’s rules, WMD value was used to reflect the clinical relevance of combination therapy: trivial < 0.2 ≤ small<0.5 ≤ medium<0.8 ≤ large.^28^ The choice of data analysis methods depended on between-study heterogeneity; it was tested by Cochran’s Q statistic and *I*^2^ statistics. When the heterogeneity test was significant (*p* ≤ 0.1, *I*^2^ ≥ 50%), a random-effects model was used; otherwise, a fixed-effects model was used. Sensitivity and subgroup analyses were performed to identify the cause of heterogeneity. A two-sided p-value lower than 0.05 indicated a significant level of difference. Publication bias was tested using a funnel plot in STATA version 14.0.

## 3 RESULTS

### 3.1 Literature search

A preliminary database search yielded 1079 papers. We removed 405 papers from them due to duplication. We then reviewed the titles and abstracts of the remaining 674 articles and excluded 632 obviously irrelevant records. After the above steps, we identified 42 pieces of literature for full-text review. In this process, 33 studies were excluded due to reasons such as improper intervention (n = 21), unclear or insufficient data (n = 5), not RCTs (n = 3), and not angina pectoris patients (n = 4). Finally, nine studies^22–24 29–34^ were included in the meta-analysis. The screening process is shown in Figure 1.

**Figure 1.**
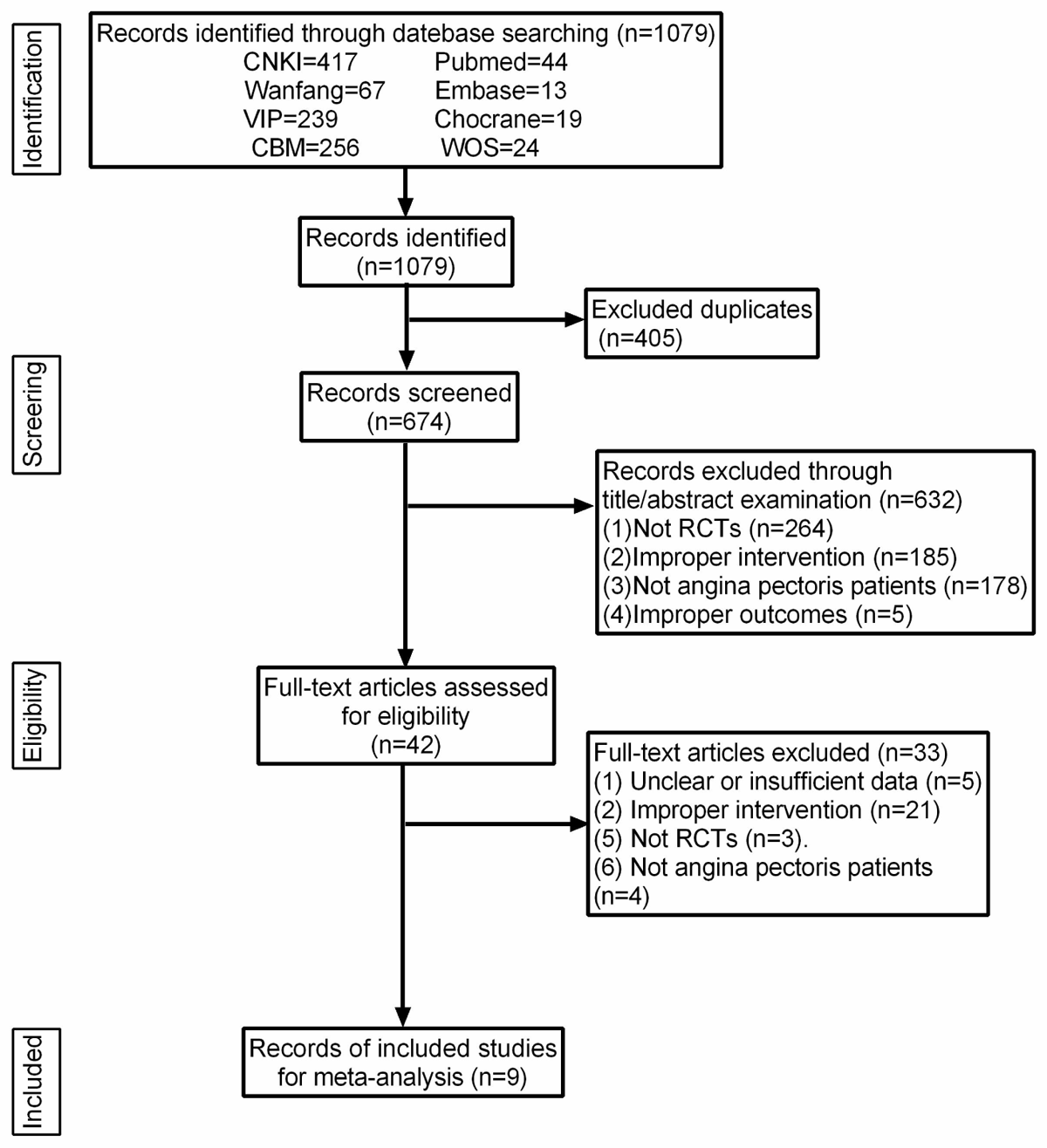
Literature search and screening process

### 3.2 Characteristics of the Studies

The final meta-analysis included nine RCT studies^22–24^ ^29–34^ conducted and published in China between 2012 and 2023. The total sample size was 754, with 378 participants in the experimental group and 376 participants in the control group. Five out of nine studies focused on stable angina patients,^22–24^ ^30^ ^31^ one study focused on unstable angina patients,^29^ ^34^ another two included all types of angina patients.^32^ ^33^ One of the studies conducted by Jia et al. was a three-arm study involving Western agency intervention (arm 1), western agency plus acupuncture intervention (arm 2), and Western agency plus acupuncture and Chinese medicine intervention (arm 3).^30^ We only included data from arms 1 and 2 for the analysis. The other eight were two-arm studies comparing acupuncture plus statin with statin alone.^22–24^ ^29^ ^31–34^

The acupuncture protocols used in each study showed differences in treatment techniques, the selection of acupoints, the length of one stimulation session, and the duration and frequency of intervention. Among the nine studies, four used manual acupuncture plus statin as the intervention treatment,^23^ ^24^ ^29^ ^32^ two used electroacupuncture plus statin,^30^ ^34^ and three used moxibustion plus statin.^22^ ^31^ ^33^ The number of acupoints applied varied across the studies. Two studies by Deng et al. and Zhao et al. used a flexible acupoint protocol, selecting the acupoints based on patients’ traditional Chinese medicine (TCM) syndrome.^29^ ^34^ The remaining seven studies used a fixed protocol, with the number of acupoints ranging from one to 12.^22–24^ ^30–33^ The stimulation time of acupuncture for most studies was from 20 to 30 minutes per session.^23–24 29 30 32 34^ However, the stimulation time per acupoint varied from 10 to 50 minutes in the remaining two studies,^22^ ^31^ depending on the duration of the de-qi sensation experienced by the patients. The duration of treatment ranged between 4 weeks and 24 weeks. As for statin, seven studies utilized atorvastatin,^23–24^ ^29–31^ ^33^ ^34^ while the other two used pravastatin and simvastatin, respectively.^22^ ^32^ For intervention frequency, six studies administered acupuncture or moxibustion treatment once a day.^22^ ^29–33^ However, in the remaining studies,^23^ ^24^ ^34^ acupuncture treatment was performed every other day or three times a week.

Seven studies included data on four lipid indicators: total cholesterol (TC), triglycerides (TG), LDL-C, and high-density lipoprotein cholesterol (HDL-C).^22^ ^29–34^ Wang (2021) focused solely on TC and TG,^24^ while Sun (2021) examined TC, TG, and LDL-C levels in patients with stable AP.^23^ Only two studies looked at AEs.^30^ ^31^ The specific characteristics of the studies included are shown in Table 2.

### 3.3 Risk of Bias

Each study included in the analysis exhibited some degree of bias. Regarding selection bias, seven studies utilized the random number table method to assign subjects.^22–24 31–34^ Therefore, we evaluated these studies as having a low risk of bias. However, two other studies were rated as unclear due to the lack of description regarding their randomization methods.^29 30^ None of the studies provided information on allocation concealment, resulting in an unclear risk in this domain for all studies. Since all the included studies were designed as open-label trials, they were all rated as having a high risk for performance bias. For detection bias, the study conducted by Wang (2021) had a low risk due to the design of blinding applied to statisticians during analyses.^24^ However, the other eight studies did not provide sufficient information on the blinding of outcome assessment.^22–23 29–34^ Therefore, these studies were rated as having an unclear risk of bias. The study protocols for all the studies were not registered in a publicly available database, which hindered us from fully knowing the protocols of each study. As a result, these studies were categorized as having an unclear risk of bias for reporting bias. We identified that the study conducted by Wang (2021) had an unclear risk of attrition bias due to seven patients dropping out for poor compliance during the study period,^24^ while other studies had a low risk in this aspect.^22–23 29–34^ All included studies were evaluated as having a low risk of other bias, as we did not find other sources of bias in each study. A summary of the risk of bias in each of the studies included is presented in Figure 2.

**Figure 2.**
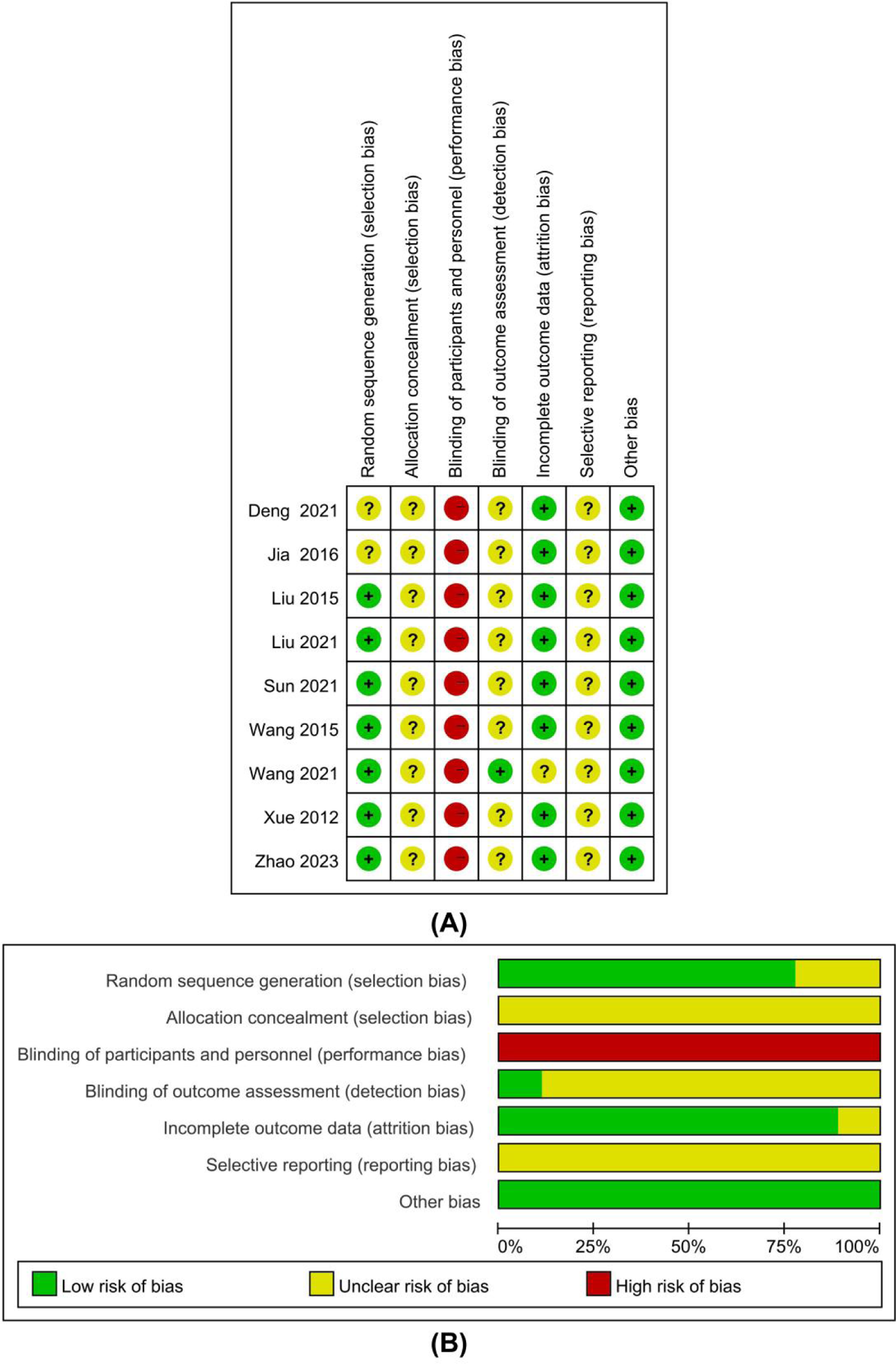
Risk of bias assessment using the Cochrane tool. (A) risk of bias summary; (B) risk of bias graph: percentage of each risk of bias item across all included studies.

### 3.4 Meta-analysis results

#### 3.4.1 Total cholesterol

Nine studies involving 754 patients investigated the impact of combining acupuncture and statin on TC levels.^22–24 29–34^ The pooled effect were summarised using a random-effect meta-analysis revealed that the combination treatment did not provide any advantages in reducing TC levels (MD=-0.75, 95% CI: −1.53 to 0.03, *P*=0.06, *I^2^*=98%) (Figure 3A). However, statistical significance was observed in this result when using a fixed effects model (MD=-1.37, 95% CI: −1.47 to −1.27, *P* < 0.00001) (online supplementary Figure 1); it indicated that the result was unstable. Subsequently, subgroup analysis showed disappeared heterogeneity and significant differences in the subgroups of moxibustion (MD=-0.53, 95% CI: −0.73 to −0.33, *P* < 0.00001, *I^2^* = 0%), electroacupuncture (MD=-0.40, 95% CI: −0.74 to −0.07, *P*=0.02, *I^2^*=0%), 4 weeks (MD=-0.47, 95% CI: −0.64 to −0.29, *P* < 0.00001, *I^2^*=0%), and ≥12 weeks (MD=-0.43, 95% CI: −0.66 to −0.20, *P*=0.0003, *I^2^*=0%). However, the manual acupuncture subgroup (MD=-1.04, 95% CI: −2.41 to 0.33, *P*=0.14, *I^2^*=99%) and 8 weeks subgroup (MD=-1.68, 95% CI: −3.45 to 0.08, *P*=0.06, *I^2^*=98%) still exhibited significant heterogeneity and no significant changes in TC levels. In sensitivity analysis, the trial from Wang (2021) was considered the source of heterogeneity. After removing this study, the overall (MD=-0.48, 95% CI: −0.61 to −0.35, *P*<0.00001) and manual acupuncture subgroup analyses (MD=-0.46, 95% CI: −0.67 to −0.25, *P*<0.0001) showed disappeared heterogeneity and significant differences (Figure 3B). We believed that too big a difference in TC levels between the groups in this trial compared to others was a vital element that resulted in the heterogeneity.

**Figure 3.**
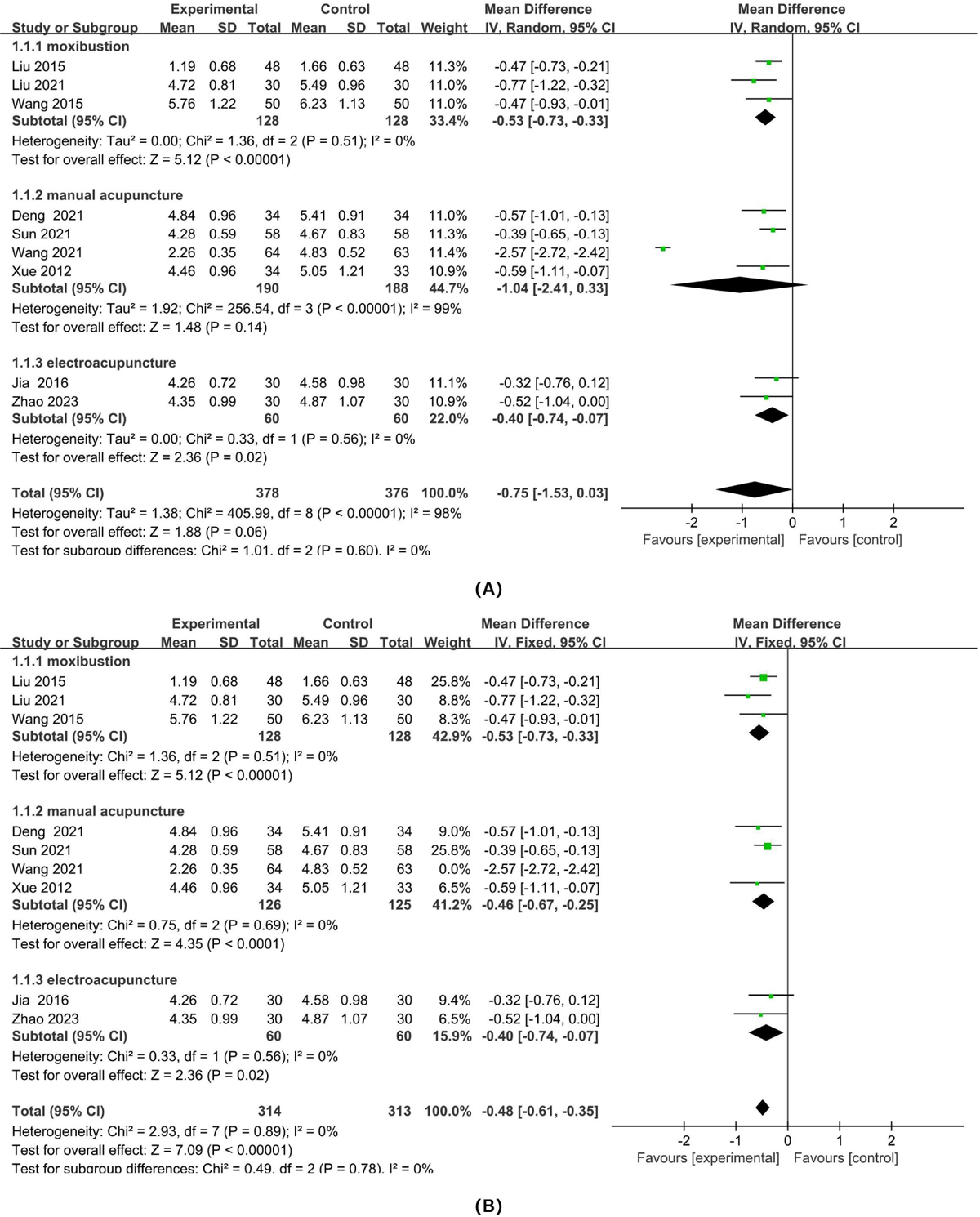
The Forest plot of subgroup analysis stratified by acupuncture types (moxibustion, manual acupuncture, and electroacupuncture) for TC levels: (A) included the trial from Wang 2021; (B) excluding the trial from Wang 2021.

#### 3.4.2 Triglycerides

Nine studies examined TG levels in patients with AP of CHD.^22–24^ ^29–34^ Due to significant heterogeneity (*I^2^*=92%), a random effect model was employed to calculate the pooled effect size of TG levels. The results indicated that the combination treatment of acupuncture and statin was more effective in reducing TG levels compared to statin alone (MD=-0.59, 95% CI: −0.86 to −0.32, *P*<0.0001) (Figure 4). This finding was consistent with both the moxibustion subgroup (MD=-0.87, 95% CI: −1.25 to −0.49, *P*<0.00001, *I^2^*=83%) and the manual acupuncture subgroup (MD=-0.54, 95% CI: −1.00 to −0.09, *P=*0.02, *I^2^*=96%) but not with the electroacupuncture subgroup (MD=-0.25, 95% CI: −0.52 to −0.02, *P=*0.07, *I^2^*=0%). Additionally, each treatment duration subgroup showed significant differences in TG levels (Table 3). The heterogeneity decreased in the electroacupuncture subgroup and each treatment duration subgroup (Figure 4, Table 3). In a sensitivity analysis, no individual study significantly influenced the overall effect size or heterogeneity, suggesting a robust result.

**Figure 4.**
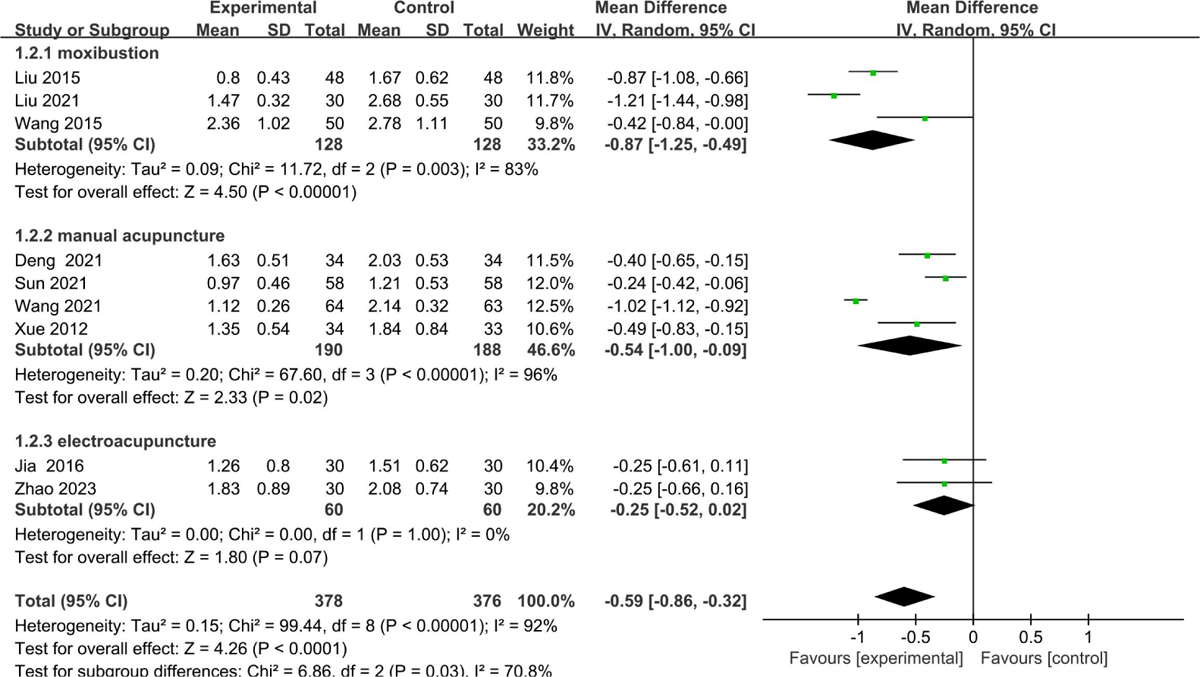
The Forest plot of subgroup analysis stratified by acupuncture types (moxibustion, manual acupuncture, and electroacupuncture) for TG levels.

**Table 3.**
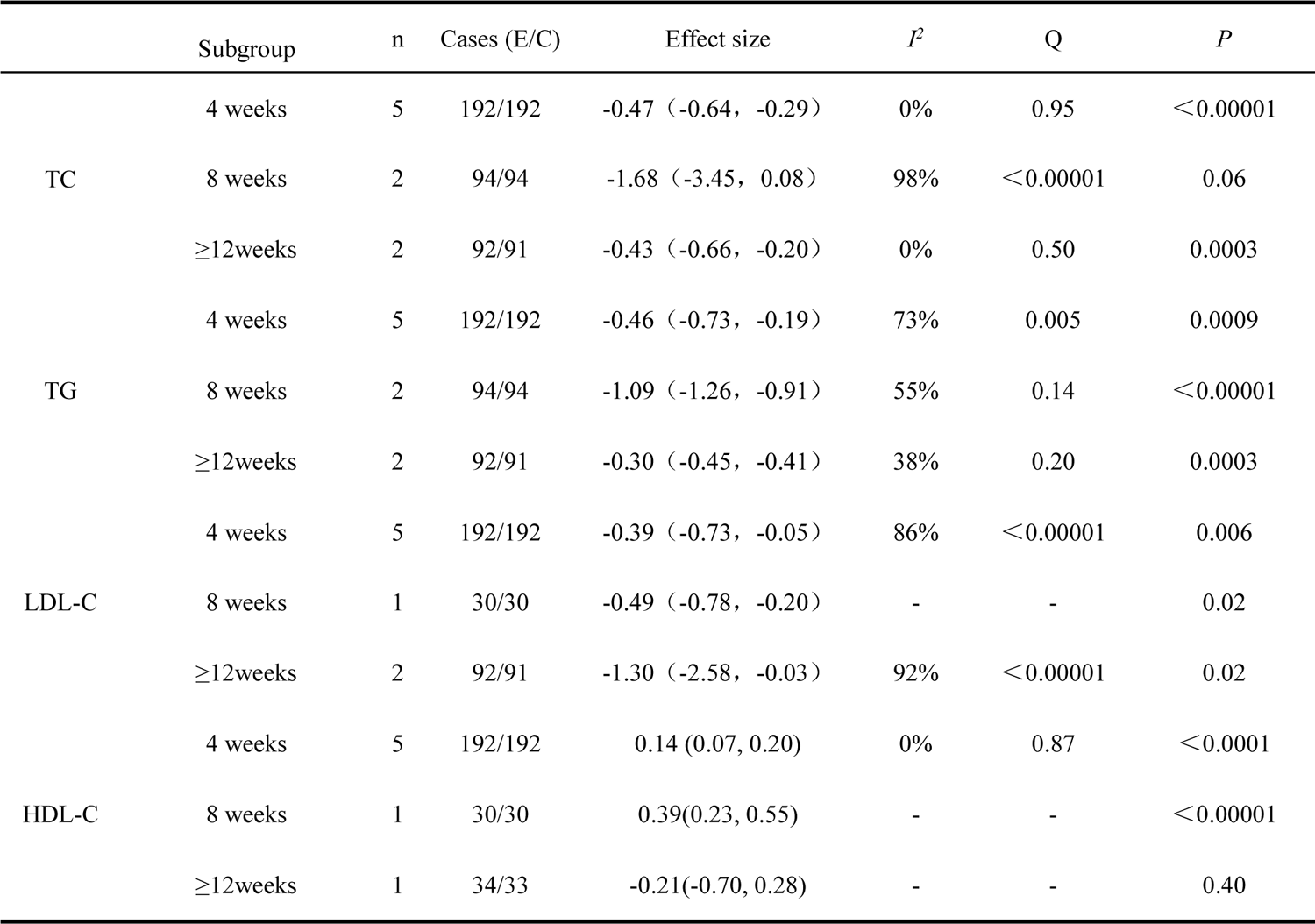
Subgroup analysis based on the duration of interventions.

#### 3.4.3 Low-density lipoprotein cholesterol

The levels of LDL-C were measured in eight studies.^22–23^ ^29–34^ As shown in Figure 5, the combined effects suggested that the combination treatment has advantages in reducing LDL-C levels (MD=-0.66, 95% CI: −0.99 to −0.33, *P*=0.0001). However, there is a significant level of heterogeneity (*I^2^*=91%) found among these studies. Subgroup analysis based on acupuncture types showed that the combination treatment was effective on LDL-C in the studies that utilized moxibustion (MD=-0.57, 95% CI: −0.98 to −0.16, *I^2^*=88%, *P*=0.006), manual acupuncture (MD=-1.00, 95% CI: −1.82 to −0.18, *I^2^*=96%, *P*=0.02), or electroacupuncture (MD=-0.30, 95% CI: −0.55 to −0.05, *I^2^*=0%, *P*=0.02). Additionally, combination treatment showed significant LDL-lowering effects regardless of whether the treatment period was 4 weeks (MD=-0.39, 95% CI: −0.73 to −0.05, *I^2^*=86%, *P*=0.03), 8 weeks (MD=-0.49, 95% CI: −0.78 to −0.20, *P*=0.001), or more than 12 weeks (MD=-1.30, 95% CI: −2.58 to −0.03, *I^2^*=97%, *P*=0.04) (Table 3). In sensitivity analysis, the final result and overall heterogeneity were not influenced by any trials.

**Figure 5.**
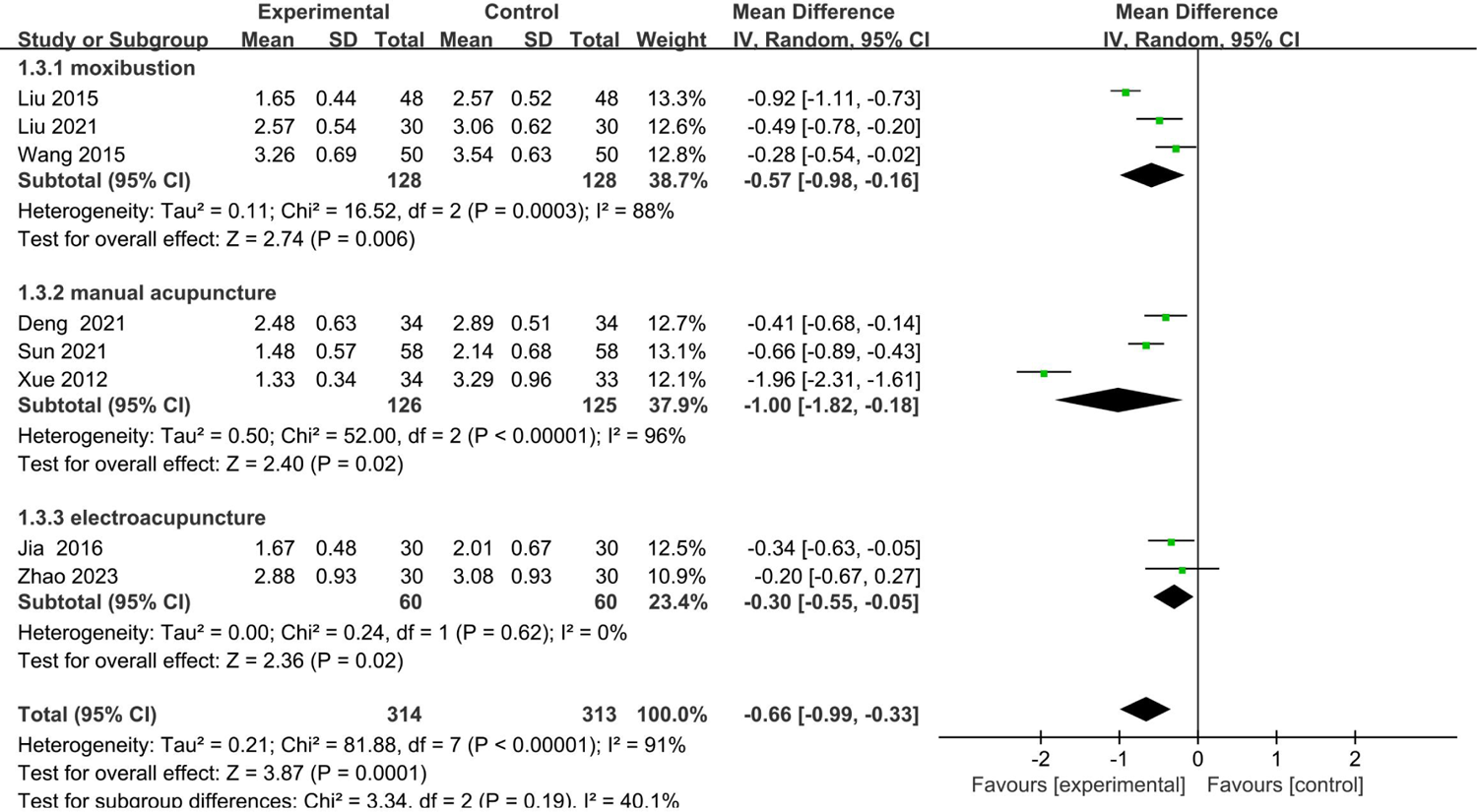
The Forest plot of subgroup analysis stratified by acupuncture types (moxibustion, manual acupuncture, and electroacupuncture) for LDL-C levels.

#### 3.4.4 High-density lipoprotein cholesterol

This meta-analysis, based on seven trials involving 501 patients with AP of CHD, demonstrated that the combining treatment of acupuncture and statin was associated with higher HDL-C levels (MD=0.16, 95% CI: 0.06 to 0.26, *P*=0.001) (Figure 6A).^22^ ^29–34^ However, moderate heterogeneity was found for this comparison (Q=0.06; *I^2^*=50%). Subgroup analysis showed a remarkable advantage when using moxibustion (MD=0.20, 95% CI: 0.04 to 0.36, *I^2^*=78%, *P*=0.02) or treatment duration lasting 4 (MD=0.14, 95% CI: 0.07 to 0.20, *I^2^*=0%, *P*<0.0001) or 8 (MD=0.39, 95% CI: 0.23 to 0.55, *P*<0.00001) weeks (Figure 6A, Table 3). However, no significant effect was observed in the subgroups of manual acupuncture (MD=0.05, 95% CI: −0.27 to 0.38, *I^2^*=50%, *P*=0.75), electroacupuncture (MD=0.11, 95% CI: −0.15 to 0.37, *P*=0.41), or ≥12 weeks (MD=-0.21, 95% CI: −0.70 to 0.28, *P*=0.40). Besides, heterogeneity reduction was observed in subgroups of electroacupuncture and 4 weeks. The sensitivity analysis suggested that the study from Liu (2021) (which had a different treatment duration compared with other trials) might contribute to the heterogeneity; the heterogeneity of the overall effect disappeared (*I^2^*=0%), and the heterogeneity of the moxibustion subgroup diminished from 78% to 5% after removing this trial (Figure 6B).

**Figure 6.**
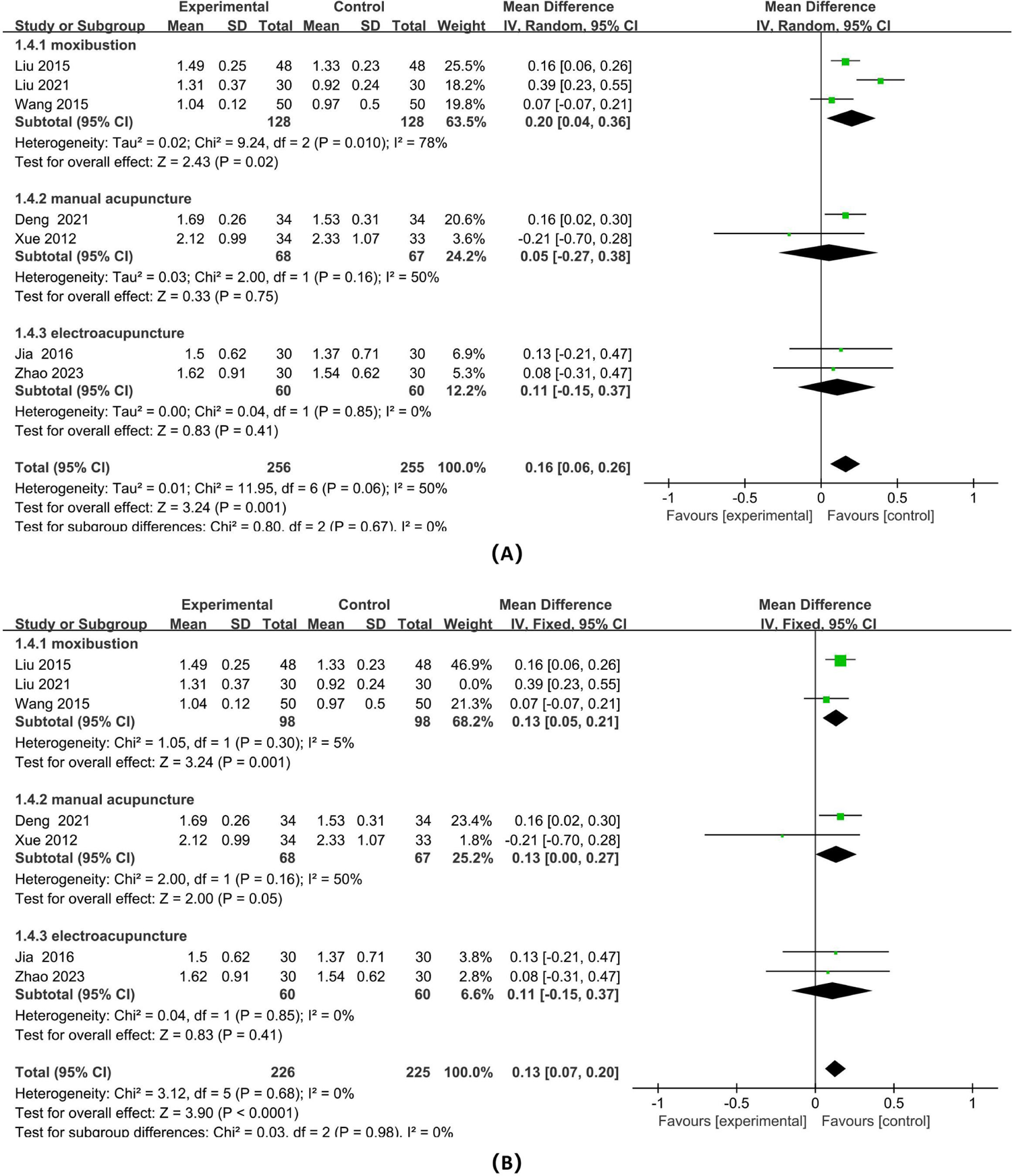
The Forest plot of subgroup analysis stratified by acupuncture types (moxibustion, manual acupuncture, and electroacupuncture) for HDL-C levels: (A) included the trial from Liu 2021; (B) excluding the trial from Liu 2021.

#### 3.4.5 Safety

Only two studies evaluated the impact of combination treatment on patients’ safety by blood indicators (blood routine, liver function, and kidney function),^30^ ^31^ electrocardiograms, and symptom observation. However, no patients in these two studies experienced adverse events.

#### 3.4.6 Publication bias

Funnel plots based on study data of TC, TG, LDL-C, and HDL-C were presented in Figure 7. The fairly asymmetric in these funnel plots indicated that these outcomes were subject to publication bias.

**Figure 7.**
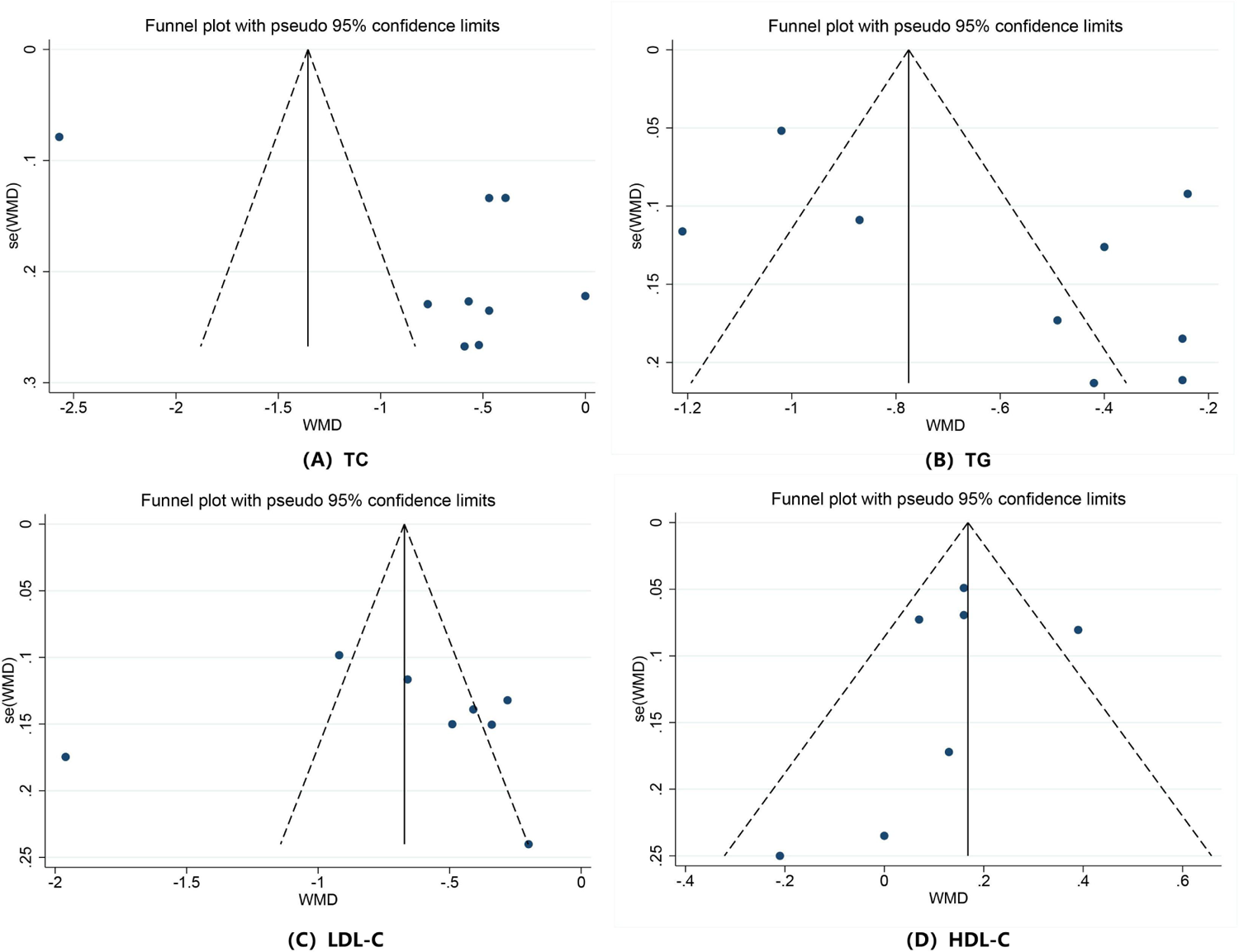
Funnel plot of the WMD versus the SE of the WMD.

#### 3.4.7 Evidence quality

According to the GRADE quality assessment criteria, the quality of evidence for the primary outcomes (TC, TG, LDL-C, and HDL-C) was assessed as ‘very low.’ We propose three primary reasons for lowering the quality of evidence. Firstly, we lowered the risk of bias by two levels for each outcome due to methodological flaws and insufficient reporting. Secondly, asymmetric funnel plots indicated the presence of publication bias in each outcome, leading us to downgrade the publication bias by one level. Thirdly, we downgraded the evidence by one level for the imprecision of each outcome as the confidence intervals were wide or crossed the line of no effect. It is worth noting that although significant heterogeneity was detected in each analysis, we did not downgrade for inconsistency as subgroup and sensitivity analyses provided reasonable explanations for the heterogeneity. Further details can be found in Table 4.

**Table 4.**
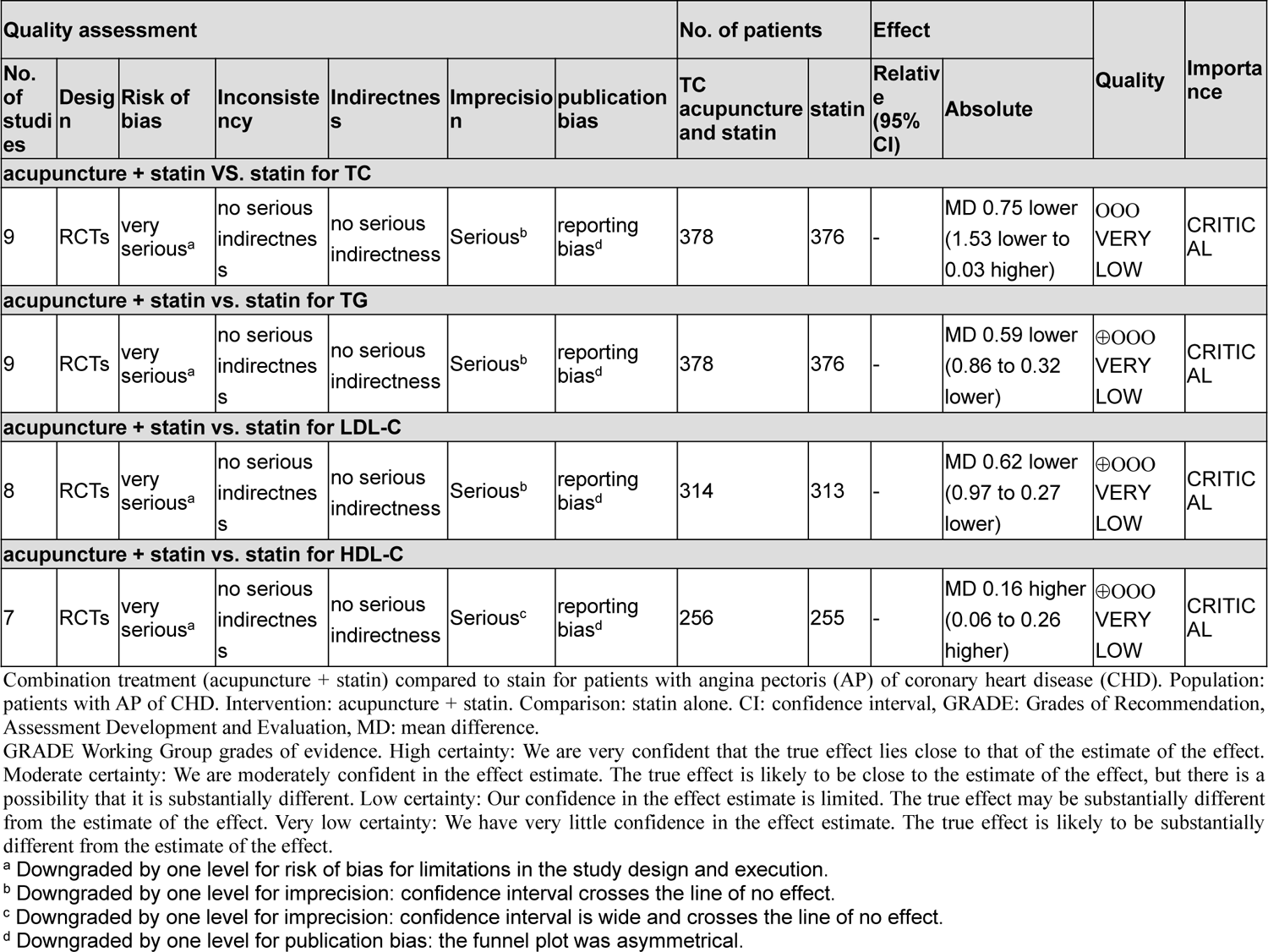
GRADE Summary of Finding Table.

| Quality assessment |  |  |  |  |  |  | No. of patients |  | Effect |  | Quality | Importance |
| --- | --- | --- | --- | --- | --- | --- | --- | --- | --- | --- | --- | --- |
| No. of studies | Design | Risk of bias | Inconsistency | Indirectness | Imprecision | publication bias | TC acupuncture and statin | statin | Relative (95% CI) | Absolute |  |  |
| acupuncture + statin VS. statin for TC |  |  |  |  |  |  |  |  |  |  |  |  |
| 9 | RCTs | very serious<sup>a</sup> | no serious indirectness | no serious indirectness | Serious<sup>b</sup> | reporting bias<sup>d</sup> | 378 | 376 | - | MD 0.75 lower (1.53 lower to 0.03 higher) | ⊖⊖⊖ VERY LOW | CRITICAL |
| acupuncture + statin vs. statin for TG |  |  |  |  |  |  |  |  |  |  |  |  |
| 9 | RCTs | very serious<sup>a</sup> | no serious indirectness | no serious indirectness | Serious<sup>b</sup> | reporting bias<sup>d</sup> | 378 | 376 | - | MD 0.59 lower (0.86 to 0.32 lower) | ⊕⊖⊖⊖ VERY LOW | CRITICAL |
| acupuncture + statin vs. statin for LDL-C |  |  |  |  |  |  |  |  |  |  |  |  |
| 8 | RCTs | very serious<sup>a</sup> | no serious indirectness | no serious indirectness | Serious<sup>b</sup> | reporting bias<sup>d</sup> | 314 | 313 | - | MD 0.62 lower (0.97 to 0.27 lower) | ⊕⊖⊖⊖ VERY LOW | CRITICAL |
| acupuncture + statin vs. statin for HDL-C |  |  |  |  |  |  |  |  |  |  |  |  |
| 7 | RCTs | very serious<sup>a</sup> | no serious indirectness | no serious indirectness | Serious<sup>c</sup> | reporting bias<sup>d</sup> | 256 | 255 | - | MD 0.16 higher (0.06 to 0.26 higher) | ⊕⊖⊖⊖ VERY LOW | CRITICAL |

Combination treatment (acupuncture + statin) compared to statin for patients with angina pectoris (AP) of coronary heart disease (CHD). Population: patients with AP of CHD. Intervention: acupuncture + statin. Comparison: statin alone. CI: confidence interval, GRADE: Grades of Recommendation, Assessment Development and Evaluation, MD: mean difference.
GRADE Working Group grades of evidence. High certainty: We are very confident that the true effect lies close to that of the estimate of the effect. Moderate certainty: We are moderately confident in the effect estimate. The true effect is likely to be close to the estimate of the effect, but there is a possibility that it is substantially different. Low certainty: Our confidence in the effect estimate is limited. The true effect may be substantially different from the estimate of the effect. Very low certainty: We have very little confidence in the effect estimate. The true effect is likely to be substantially different from the estimate of the effect.
<sup>a</sup> Downgraded by one level for risk of bias for limitations in the study design and execution.
<sup>b</sup> Downgraded by one level for imprecision: confidence interval crosses the line of no effect.
<sup>c</sup> Downgraded by one level for imprecision: confidence interval is wide and crosses the line of no effect.
<sup>d</sup> Downgraded by one level for publication bias: the funnel plot was asymmetrical.

## 4 Discussion

This study is the first to investigate the effects of combining acupuncture with statin therapy on lipid levels in patients with AP of CHD. The comprehensive findings of this review demonstrated that the combination treatment has a synergistic effect and is more effective in improving TC, TG, LDL-C, and HDL-C levels compared to statin alone. It is important to note that only two studies examined AEs and provided limited data on the safety of the combination treatment. However, the results of TC levels were not robust, and evidence quality for each primary outcome was very low, so it is necessary to interpret these conclusions cautiously.

Acupuncture, a non-pharmacological intervention founded in Traditional Chinese Medicine (TCM), has proven to protect the cardiovascular system by regulating vascular tension,^35^ improving myocardial damage,^36^ reversing endothelial dysfunction,^37^ and regulating lipid metabolism.^20^ As an adjuvant therapy, acupuncture has been applied to treat CHD for many years. Several meta-analyses have examined the effectiveness and safety of acupuncture for AP of CHD.^16^ ^17^ ^18^ However, most of these studies have primarily focused on the impact of acupuncture on angina symptoms, with only a few assessing its effect on lipid levels. One such analysis, published in 2015, conducted by Zhang et al., indicated that acupuncture treatment led to a better reduction in TG and LDL-C and an increase in HDL-C levels but no change in TC levels.^38^ They did not focus on the effects of acupuncture combined with statin and included only 3 trials comparing acupuncture treatment alone to waitlist or Chinese medicine. Zhou’s study also provides supporting evidence that acupuncture helps regulate lipid metabolism in patients with AP of CHD.^39^ However, they collected three articles evaluating the combined therapy of acupuncture and Chinese medicine to conduct their analysis, which makes it difficult to separate the effect of acupuncture from Chinese medicine. Compared to the two mentioned studies, the present review has several advantages. Firstly, we restricted the eligible intervention to acupuncture combined with statin versus statin, ensuring the comparability among the included studies. Secondly, we assembled a relatively large number of studies (n=9) in this systematic review, which provided more objective and sufficient data to investigate the association between combined therapy (acupuncture + statin) and lipid levels. Thirdly, we carefully considered the effect of heterogeneity and conducted subgroup analyses to identify relevant heterogeneity moderators. Additionally, we performed a sensitivity analysis to assess the impact of each study on the overall effect size and heterogeneity.

Despite combination treatment showing a significant effect in regulating lipid levels, heterogeneity was detected in this review. The differences in acupuncture protocols among the studies bring important ideas to explore the heterogeneity. The subgroup analyses based on the types of acupuncture and duration of intervention resulted in attenuation in the overall heterogeneity for each primary outcome, indicating between-study heterogeneity may be partly derived from these factors. Then, we also found remarkable diversity from acupoint selection across included studies, which may also be the cause of heterogeneity. However, the included studies were too heterogeneous and sparse, which prevented us from analyzing the impact of acupoints on heterogeneity by a further subgroup analysis. Moreover, the types of acupuncture have also been suggested to be factors that determined the effectiveness of combination treatment in regulating TG and HDL-C levels. Subgroup analyses showed that combination treatment reduction of TG levels was associated with moxibustion and manual acupuncture but not electroacupuncture. Additionally, HDL-C levels elevated when statins were combined with moxibustion, not manual acupuncture or electroacupuncture, suggesting that moxibustion, the non-invasive warm stimulation, may be more suitable for regulating HDL-C levels compared with an acupuncture needle, the invasive mechanical stimulation. However, the MD values (MD = 0.20) in the moxibustion subgroup for HDL-C levels were low, revealing questionable clinical relevance. Moreover, the subgroup analyses also showed that combination treatment did not further elevate the HDL-C levels over ≥12 weeks of treatment duration. However, this outcome was based on data from one single trial,^32^ having a chanciness. Further research is required to explore the relationship between the hypolipidemic effect of acupuncture and factors such as acupuncture type, intervention duration, and acupoints. Additionally, determining the best acupuncture treatment plan for lipid-lowering is necessary to improve the consistency and reliability of future research.

In the sensitivity analysis of the TC levels, we observed a significant effect and disappeared heterogeneity after eliminating the trial from Wang (2021);^24^ thus, this trial was accountable for the unstable results and the substantial heterogeneity. In the study by Wang (2021), a too big difference in TC levels between the groups may be the cause that affects the heterogeneity and overall effect size. For TG and LDL-C levels, the direction of results and levels of heterogeneity were maintained after removing each single study one by one; thus, these results were stable. For HDL-C levels, the study by Liu (2021)^31^ is considered responsible for the high heterogeneity, as the heterogeneity of the overall effect was reduced from 50% to 0% when this study was removed. Using an 8-week intervention differing from other trials in this trial may be the reason for heterogeneity.

Although our research suggests that acupuncture provides additional reduction in lipid levels in patients with AP of CHD when added to statin therapy, this finding is insufficient to inform clinical practice. Firstly, LDL-C has always been considered a primary indicator for evaluating cardiovascular risk.^7^ The latest Chinese guidelines for lipid management recommend that the LDL-C goal for patients at extremely high cardiovascular disease risk is < 1.8mmol/L and a ≥50% LDL-C reduction.^14^ In this review, all included studies were conducted in a Chinese population. However, these studies used lipid metabolism indicators as secondary measures, focusing only on the mean difference in blood lipid levels within and between groups without calculating the proportion of patients achieving lipid goals in each group. Therefore, the current research does not fully capture the clinical significance of combining acupuncture and statin treatment. Secondly, evidence shows that, when combined with chemical drugs (such as anesthetic,^40^ hormone therapy,^41^ and fluoxetine^42^), acupuncture can improve the effectiveness, meanwhile reducing the occurrence of AEs of drugs, enhancing their safety. Thus, we believe that acupuncture with this advantage is promising to provide more clinical benefit for statin-intolerant patients. However, only two included studies investigated the AEs,^30^ ^31^ and no AEs occurred in either study. Such limited data make it difficult to validly evaluate the impact of acupuncture add-on treatment on the safety of statin use. Additionally, clearing the impact of acupuncture plus statin treatment on the risk of major adverse cardiovascular events (MACE) in patients with AP of CHD can provide more reference for the clinical application of combined therapy of acupuncture and statin. However, all included studies have not focused on this issue. Therefore, in the future, more well-design clinical studies that focus on acupuncture combined with a statin for lipid levels in patients with AP of CHD should be carried out and should use blood lipids as primary observation indicators and pay more attention to safety and the risk of MACE to comprehensively examine the actual clinical benefit of lipid-lowering effect by combination treatment for patients with AP of CHD.

The meta-analysis has several limitations that should be taken into account. Firstly, all the studies included in this review had open-label study designs and lacked detailed descriptions of allocation concealment and evaluator blindness. This increases the risk of bias across the included studies, which is an important factor in downgrading the quality of evidence. Secondly, lifestyle, diet, and exercise are underlying factors that influence the lipid levels but are not reported or assessed in any of the included studies. Thirdly, none of the included studies registered their protocols online. Fourthly, it is important to note that all the eligible studies were conducted in China, which may limit the generalizability of the findings to other parts of the world. Fifthly, each included study in this review is a single-center design with a small sample size. Finally, the endpoints analyzed in this review were limited to TC, TG, LDL-C, and HDL-C, without considering other key indicators of lipid metabolism such as apoB, apoA, very LDL-C, small dense LDL-C, and TG/HDL-C, which are important for predicting cardiovascular events. Given these limitations, caution should be exercised when interpreting the results. Future research should address these shortcomings and strive to improve them to provide more high-quality evidence and validate the reliability of the findings.

## 5 Conclusions

Our findings revealed that combining acupuncture with statin treatment resulted in a more significant improvement in lipid levels, such as TC, TG, LDL-C, and HDL-C, in patients with AP of CHD compared to statin treatment alone. These results suggest that acupuncture could be a promising adjunctive approach for lipid management. However, drawing general conclusions from unstable and very low-quality outcomes is unwise. Therefore, while this review added new insights into the treatment of AP of CHD with acupuncture, the effect and safety of acupuncture combined with statin for lipid levels should be further confirmed by more high-quality research.

## Contributions

Xiao-Fang Yang and Mi Liu developed the foundational concepts for this review. Zhao-Bo Yan and Xian-Ming Wu conducted the literature search and data extraction, while Zhi-Hong Yang and Ning Zhang were responsible for data analysis; Zhao-Bo Yan and Mai-Lan Liu were responsible for drafting the manuscript, while Jiao-Jiao Xiong assisted with the illustrations. Xiao-Fang Yang and Mi Liu revised the manuscript. All authors contributed to the writing of the manuscript and approved the final version.

## Patient and public involvement

Patients and/or the public were not involved in the design, or conduct, or reporting or dissemination plans of this research.

## Patient consent for publication

Not applicable.

## Ethics approval

Not applicable.

## Provenance and peer review

Not commissioned; externally peer reviewed.

## Data Availability

All data produced in the present work are contained in the manuscript.

---

**Supplementary table 1.**
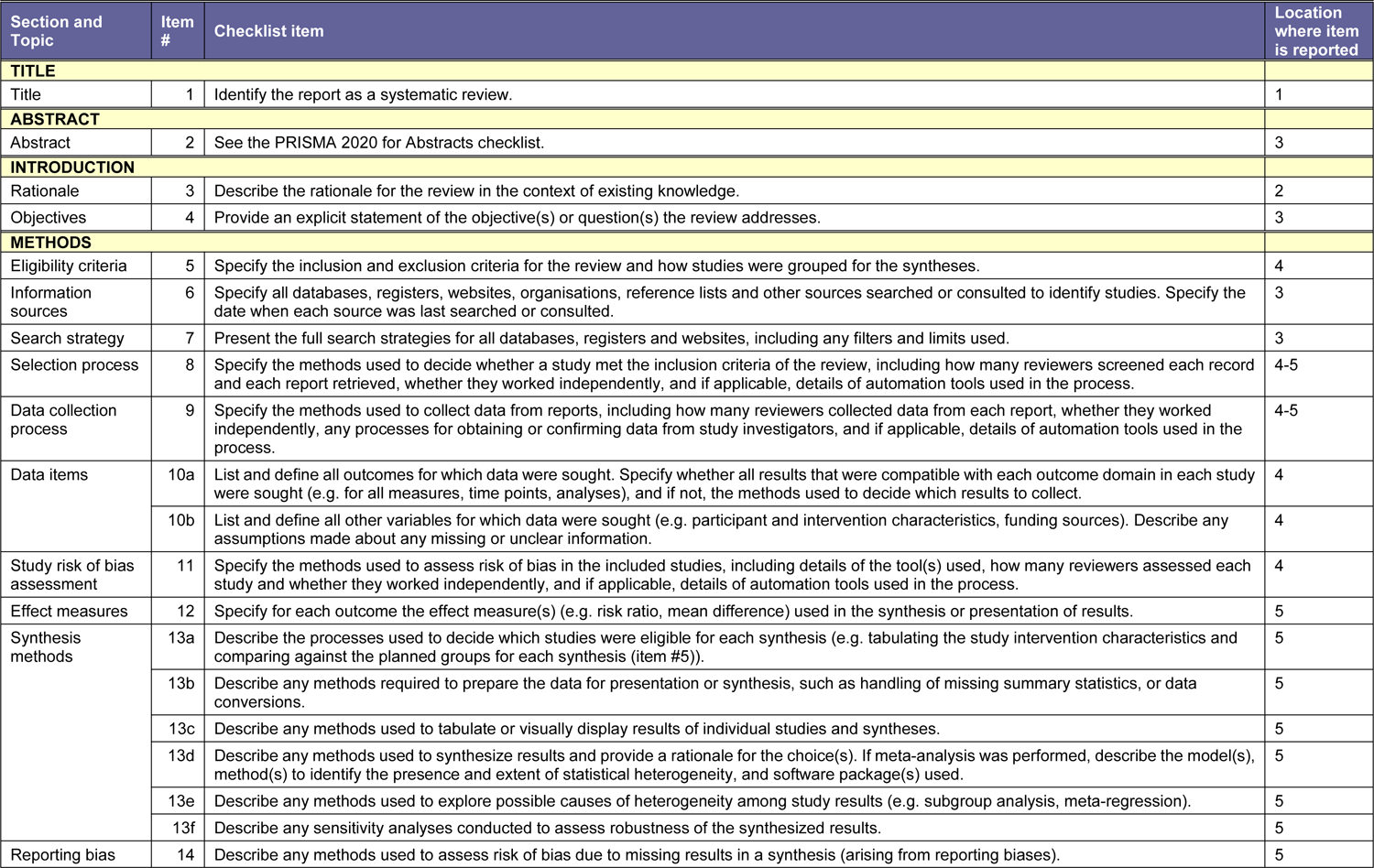
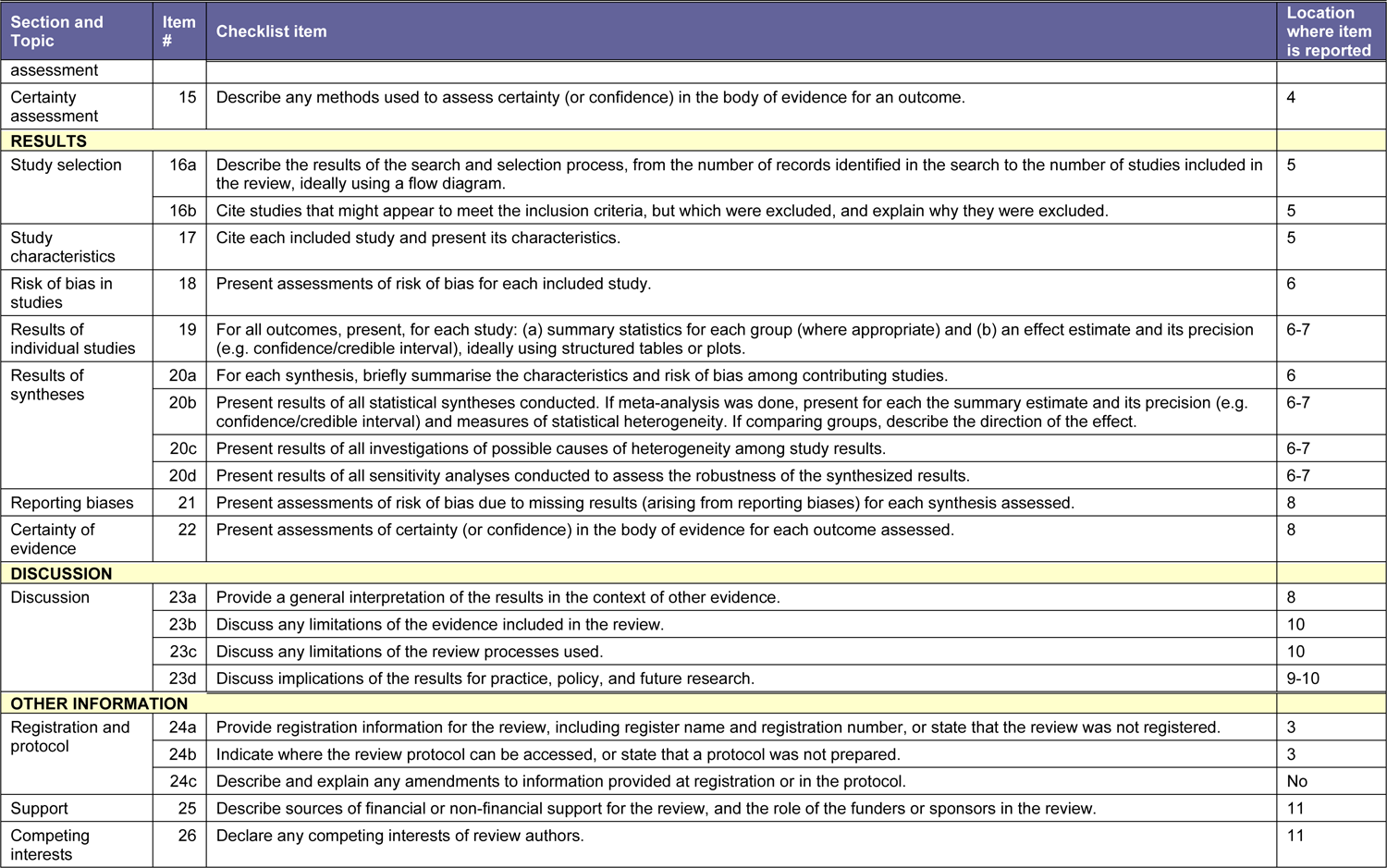
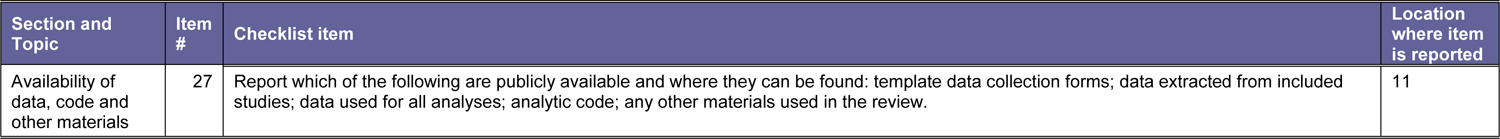
Details of the PRISMA checklist.

**Supplementary figure 1.**
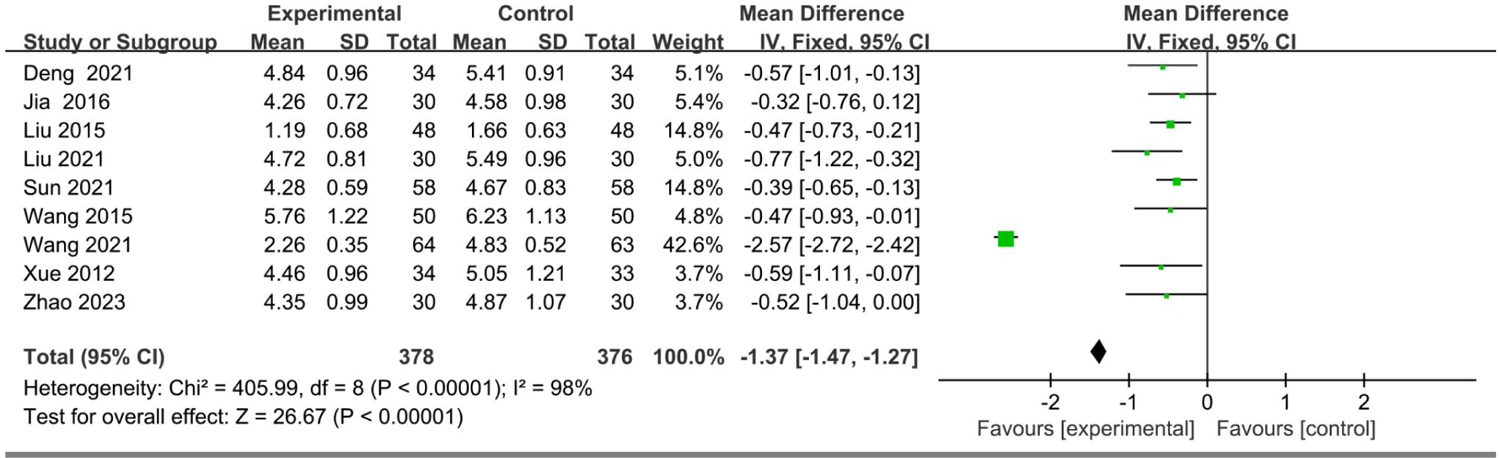
The forest plot showing mean difference and 95% confidence intervals (CIs) for the effect of combination treatment (acupuncture + statin) on TC levels.

